# A protocol for a critical realist systematic synthesis of interventions to promote pupils’ wellbeing by improving the school climate in Low- and Middle-Income Countries

**DOI:** 10.1101/2023.05.18.23290176

**Authors:** Pamela Abbott, Rachel Shanks, Isabel Stanley, Lucia D’Ambruoso

## Abstract

**Introduction:** The review described in this protocol will be the first critical realist review of the literature reporting on the impact of interventions to promote pupils’ wellbeing by improving the school climate in Low- and Middle-Income Countries. The review is being carried out to inform the programme theory for a critical realist evaluation of a whole school mindfulness intervention in Ethiopia and Rwanda to improve pupils’ mental wellbeing. Our initial programme theory hypothesises that pupils’ (and teachers’) responses to the mindfulness intervention as well as changing the behaviour and attitudes of individual pupils and teachers, will change the ‘school climate’ in ways that have a positive impact on mental wellbeing. This literature review will facilitate the identification of mechanisms for change working at the level of the whole school climate, something which is only infrequently discussed in evaluations of mindfulness interventions.

**Methods and analysis:** A critical realist review methodology will be used to provide a causal interdisciplinary understanding of how school climate can promote the wellbeing of pupils. This will be done through a systematic literature review and extrapolating context, agency, intervention, mechanisms, and outcome configurations and synthesising these to provide a conceptual understanding of the impact of interventions to improve school climate.

**Discussion:** The review findings will inform a critical realist evaluation of a mindfulness intervention in schools that we will be carrying out. The findings from the review will enable us to focus more precisely and transparently on what policymakers and other stakeholders need to know about how school climate changes due to introducing mindfulness to the curriculum and how this impacts pupils’ wellbeing [and for which pupils]. We will publish the findings from the review in academic and professional publications, policy briefs, workshops, conferences, and social media.

**PROSPERO registration number:** CRD42023417735

## Introduction

The review described in this protocol will be the first systematic critical realist review of the literature reporting on the impact of the ‘school climate’ (defined here as schools’ structural, interpersonal relations and teaching practices, and cultural norms and values) on pupils’ wellbeing in Low- and Middle-Income Countries (LMICs). The review is being carried out to inform the programme theory for a critical realist evaluation of a whole school-based mindfulness intervention (SBMI) in Ethiopia and Rwanda on pupils’ mental wellbeing (1,2). Our initial programme theory hypothesises that pupils (and teachers) will be able to use the psychological resources they gain through the mindfulness intervention to change the ‘school climate’ in ways that positively impact pupils’ mental wellbeing.

Our review is novel. Existing literature on the impact of mindfulness interventions focuses on individual psychological outcomes. It rarely considers the pathways to improved mental wellbeing through changes in the social structural or cultural contexts due to changes in the behaviour of pupils and teachers wrought by mindfulness interventions (3–5). This literature review will yield a framework of existing theories as a guide to identifying and understanding the underlying process (mechanisms) that shape the ‘school climate’ and identify those that may be triggered by a whole school mindfulness intervention and promote pupils’ mental wellbeing.

Critical realists recognise that not every intervention will work for each person in the same way or different contexts in the same way. While traditional reviews have been concerned with descriptive outcomes and average effects, critical realists are more concerned with exploring how interventions work, for whom and under what circumstances. The broad purpose of the review is to move from empirical observation to develop a theorised understanding of the impact of school climate on pupils’ wellbeing in LMICs and identify aspects of school climate that can be linked to mechanisms triggered by mindfulness interventions. This will produce knowledge that enables us to make recommendations to improve practice, that is, improve the school climate and deliver mindfulness interventions in schools (6). The main purpose is to build a middle-range theory (7) that models the underlying mechanisms influencing the school climate. We will use the critical realist RRRIREI© (resolution, redescription, retroduction, retrodiction, elimination, identification, correction) framework for explanatory interdisciplinary research (8,9). The findings will enable us to refine the programme theory for research we are carrying out examining the potential for school-based mindfulness interventions to promote the mental wellbeing of children and adolescents in Rwanda and Ethiopia. The findings will also support policymakers and others to implement policies to improve the school climate and promote the wellbeing of children and adolescents in LMICs more generally by providing an understanding of under what conditions and for which pupils’ interventions make the school climate more positive.

The quality of school life (the school climate) is determined by a combination of the structural, interpersonal relations and teaching practices, and the cultural norms and values of the school as influenced by the wider social (‘laminated’) system (Table 1) in which it is embedded (10). It is crucial for promoting safer, more supportive and more civil schools (11). School climate improvement measures make schools friendlier, pupils (and teachers) more connected to the school and prevent dropout (11–14). Four sub-constructs of the school climate have been identified in the literature that impacts pupils’ wellbeing: safety (feeling safe in school), relationships (e.g. school connectedness/engagement, social support, leadership and pupils’ perception of the school climate), teaching and learning (academic environment) and the institutional environment (school connectedness) (11–14). The authors argue that these four sub-constructs are interconnected, influencing each other and being influenced by the broader context (laminated system) in which schools are located.

**Table 1:** Critical Realist Terminology

|  |  |
| --- | --- |
| Depth Ontology | The surface-level events and experiences we perceive (the empirical) are manifestations of underlying causal powers, |
|  | structures, and mechanisms. These deeper aspects are not directly observable but have a causal influence on the observable phenomena. |
| Complexity | The social world is dynamic, non-linear, and has emergent properties giving rise to the diverse and multifaceted phenomena we observe. |
| Demi-regularities | Demi-regularities capture the idea that there are patterns in the world that tend to occur under certain conditions or contexts, but these regularities are not fixed or invariant and are sensitive to contextual factors. |
| Judgemental Rationalism | Making claims to knowledge and truth by determining the theory with the greatest explanatory power. |
| Laminated System | The social world comprises distinct layers or dimensions from the physical, chemical and biological to the global system. These layers are interconnected and mutually constitutive, meaning that changes or processes at one level can have consequences for other levels. Laminated systems have emergent properties; that is, the complex interactions and relationships between the layers of a laminated system give rise to new mechanisms that cannot be reduced to their component parts. |
| Mechanisms | The underlying causal powers or processes that generate and shape observable phenomena in the social and natural world. They are not directly observable but are inferred from the patterns and regularities observed in the empirical data. Contextual mechanisms are inherent to the system and arise from the complex interactions and relationships within the system. Intervention mechanisms are interventions that can disrupt or modify a system. |
| Middle Range Theories | A middle-range theory bridges the gap between abstract, general theories and specific, concrete empirical observations. Middle-range theories typically propose causal relationships or mechanisms that explain how certain variables or factors interact to produce specific outcomes or behaviours. |
| Retrodiction | Using theoretical concepts and causal mechanisms to develop explanatory frameworks. |
| Retroduction | A form of inference that aims to uncover the underlying causal mechanisms or structures that give rise to observed phenomena. It goes beyond observing regular patterns or correlations and seeks to identify the generative mechanisms that produce those patterns. |

Schools contain both risk and protective factors for pupils’ wellbeing. An extensive body of evidence, mainly from research carried out in the global North, shows that the school climate impacts pupils’ and their teachers’ wellbeing and that there is an association between a positive school climate and wellbeing (11,12,14–29). A positive school climate influences academic outcomes (11,22–24), reduces the rates of violence in schools (11,21,22,25,26), supports skills development (21,27), promotes wellbeing and reduces the risk of mental health disorders (11,12,21,28,29). Systematic reviews have found the strongest association between pupils’ psychological wellbeing, mental health and risk behaviours on the one hand and the quality of their relationships with their fellow pupils and teachers on the other (12,14,17,19,22). Bullying and harassment, particularly, are associated with poor wellbeing and low educational achievement. A positive school climate also promotes teachers’ wellbeing and improves their relationship with pupils, which positively impacts pupils’ wellbeing (16).

While much of the research is non-experimental and cross-sectional, experimental and longitudinal research has found that the school-level socio-educational environments at baseline predict students’ wellbeing at three years follow-ups. Student perceptions of the socio-educational environment also predict their wellbeing (15,29). However, it should be noted that there is possibly a publication bias as the findings from research on interventions that have not led to improvements in the school climate are less likely to be published.

Nevertheless, the evidence is sufficient to indicate an established ‘demi-regularity’ that there is, at least in some contexts and for some pupils, an association between an intervention designed to make the school climate more positive and improving pupils’ wellbeing. It is this that needs to be explained (30).

The evidence base for a relationship between school climate and student wellbeing outcomes in LMICs is more limited. A recent (end date of search January 2019) systematic review of the association between school climate and socio-cultural, behavioural, and academic outcomes in LMICs found 35 peer-reviewed articles that met their inclusion criteria (20). All but two of the included studies reported a positive association between an intervention to improve the school climate and positive outcomes, with similar associations found as in research in high-income countries (HICs). More specifically, the findings from the review suggest a relationship between a negative school climate and bullying and violence and a positive school climate and academic achievement and wellbeing. Only seven studies included a low-income country.

However, few studies address the question of how school-based climate interventions work. Systematic reviews (and the studies included in the reviews) examine the empirical evidence on the health, behavioural and attainment effects of school environments. Still, they do not usually consider what theories might explain those changes or how they came about. One systematic review of theories of how school-based climate interventions (SBCIs) work built a comprehensive interdisciplinary theory of school effects drawing on 24 theories that partially explain the pathway from SBCIs to impact. This enabled the authors to integrate upstream, medical and downstream aspects of causal pathways (31) and test the interdisciplinary theory they built in a randomised control trial (32). The integrated theoretical model takes account of complexity and feedback loops and theorises that school climate influences pupils at multiple interacting levels: student-school commitment, student peer-commitment, student cognition and student behaviours. Imperfect measures constrained the trial. A low teacher response rate, the positive impact on pupils’ health outcomes and positive views on the changes in the school climate only became evident at the end of the trial. Nevertheless, the findings suggest that observed positive changes in pupils’ health, including mental health were the outcome of modifying the school climate.

## Aim and Objectives

This review will build on previous research on the relationship between school climate and pupil wellbeing by:

1. Including all documents from LMICs that can help with answering our research questions, including the grey literature, reports of reviews of research and discussions of theories relevant to the impact of school climate on pupil’s wellbeing;
2. Using a critical realist methodology to build a theory of what makes SBCIs work, how, where, with whom, and to what extent, focusing on LMICs.
3. Identifying context and intervention mechanisms to inform the programme theory of change for SBMI designed to promote the mental wellbeing of children and adolescents in Rwanda and Ethiopia.

The aims of the review are to:

a. describe plausible explanations for the effectiveness of SBCIs designed to promote pupils’ wellbeing in LMICs;
b. create transferable theories that can inform programme design and implementation in different settings;
c. use the findings together with our findings from systematic reviews of mindfulness interventions and theories explaining how SBMIs impact pupils’ mental wellbeing to refine our initial programme theory of how mindfulness interventions work to promote pupils’ mental wellbeing.

To achieve the aims, the objectives are to identify:

- theories about how SBCIs work in schools;
- the contexts and mechanisms that may facilitate or hinder implementation;
- how pupils and teachers respond to SBCIs (agency);
- how school contexts (social, structural, and cultural) influence the agency of pupils in responding to SBCIs and trigger mechanisms that change the context and lead to positive outcomes;
- how the school system changes (roles and relationships), including pupil-teacher relations and pupil-pupil-relations;
- how school attitudes and values change (culture), and;
- the outcomes resulting from the interventions.

## Methodology

### Introduction

To achieve our aims and objectives, we will conduct a systematic critical realist synthesis review to identify how SBCIs promote pupils’ wellbeing. A critical realist review is explanatory; it seeks to explain how interventions work and generate different outcomes in different contexts (33,34). We will explore how SBCIs are supported or inhibited by contextual mechanisms in schools, how pupils and teachers respond to them and the outcomes that result from the interaction between the intervention and contextual mechanisms and the response of pupils and teachers. In doing so, we will identify the ‘demi-regularities’, the contexts over time and space in which SBCIs enable pupils’ agency to trigger mechanisms that promote their wellbeing and improve their attainment (30). Because critical realism affirms the reality of objects, agents, and mechanisms, which cannot be viewed directly but only deduced from their effects, it seeks to identify, by retroduction and retrodiction, and judgemental rationalism, the middle-range interdisciplinary theory(s) (Table 1) that most comprehensively explain how SBCIs work. Such theories are always open to refinement in the light of new evidence.

There is no agreed standard or guide for critical realist reviews. However, a critical realist meta-theory underpins them, and they draw on some elements of the realist review methodology of Roy Pawson (6). We have adapted the standard for realist reviews (35,36) and taken into account recommendations for traditional systematic reviews (37–39).

PRISMA offers transparency, validity, replicability, and updateability (Additional Material 1).

### A critical realist research paradigm

Our methodology is informed by critical realist meta-theory. It has informed the development of methods for systematic reviews and impact evaluations that are designed to explain how and why interventions work in the ways they do (40–43). It also provides guidelines for inter/transdisciplinary research (9,44–46), including research on promoting wellbeing (9,47).

Wellbeing is a bio-psycho-social phenomenon, and the outcomes of SBCIs are likely due to the complex interaction of biological, psychological, social-structural and cultural mechanisms and require the development of middle-range theories that integrate theories from these disciplines (47–50).

The main elements of critical realism are: a depth ontology, that society is real but is only knowable through its effects; a relativist epistemology, that our understanding of the real is always partial and open to refinement or refutation; rational judgment is used to determine what explanations (social theories) are most plausible; that structure and agency are both important; that people are shaped by the context in which they live but that they can change it through their agency; that the social world is an open system and complex; and a commitment to social justice, to improve the lives of people.

Critical realism has influenced a number of approaches to carrying out social and health research, and our approach draws on Margaret Archer’s practical morphogenic approach (51,52). This methodology complements critical realism’s social ontology (53). She argues that every theory about the social involves understanding the relationship between structure, agency, and culture. The context in which agents live shapes their beliefs, desires, and opportunities and limits their agency – contextual conditioning. However, the interaction between context and agency shapes and reshapes the context; agency can change the context (morphogenesis) or reproduce it (morphostasis). An intervention designed to improve the school climate gives pupils and teachers the resources to trigger mechanisms that can lead to material and cultural change. Their responses to the intervention are shaped, but not determined, by the context, and individuals generate outcomes through actions and interactions. When actors trigger new mechanisms (material and/or cultural), the context changes. However, pupils and teachers can resist, redefine, repudiate, suspend, or circumvent engagement with the intervention.

## Methods/Design

### Step 1: establishing the scope of the work

The review will focus on SBCIs designed to promote the wellbeing of pupils in LMICs. We will document differences between SBCIs underpinned by different theories, aims, approaches and techniques and delivered differently for various lengths of time (54,55). The review will also capture other individual differences and programme characteristics that can affect programme reception by pupils and teachers and impact (56). We will only include documents relevant to the relationship between pupils’ academic achievement, behavioural, cognitive, and mental wellbeing and outcomes. Nevertheless, outcomes will likely vary by the precise nature of the SBCIs.

### Step 2: search for evidence

#### Search techniques

A rigorous systematic Preferred Reporting Items for Systematic Reviews and Meta-Analysis (PRISMA) approach will be used to search for literature (57). A PRISMA diagram will show the steps of the inclusion and exclusion of documents (Additional Material 2). The literature search will be in three phases: searching electronic databases, searching other sources such as relevant journals and core publishers, and citation tracking to ensure all relevant studies are included. The aim is to include as wide a range as possible of academic and grey literature without restrictions on study type or publication date. Literature from all LMICs will be included. Databases that index health, psychology, sociology and/or education literature will be searched. The search will be restricted to publications in the English language. The search terms and the databases used are based on the advice of an academic librarian.

The Search terms will be:

‘school’ or ‘educational context’ and ‘ethos’ or’ environment’ or ‘culture’ or ‘governance’ or ‘context’ or ‘climate’ or ‘structure’ or ‘relations’ or ‘relationships’ and ‘children’ or ‘adolescents’ or ‘youth’ or ‘young people’ or ‘juvenile’ or ‘teen’ or ‘young adult’ or ‘teenager’ or ‘pupils’ and ‘wellbeing’ or ‘mental health’ or ‘resilience’ or ‘attainment’ or’ school grades’ or ‘motivation’ or ‘connectedness’ or ‘engagement’ or ‘suicidal behaviour’ or ‘depression’ or ‘suicidal ideation’ or ‘prosocial behaviour’ or ‘risk’ or ‘risk behaviour’ or ‘burnout’ or ‘school adjustment’ or ‘attitudes’ or ‘psychosomatic complaints’ or ‘Post-traumatic stress disorder’ or ‘life satisfaction’ or ‘quality of life’ or ‘emotional’ or ‘communication’ or ‘supportive’ or ‘support’ or ‘caring’ or ‘respect’ or ‘belonging’ or’ quality of education’ or ‘anxiety’ or ‘conflict’ or ‘conduct’ or ‘bullying’ or ‘harassment’ or’ violence’ or ‘aggression’ or ‘corporal punishment’ or ‘discipline’ or ‘disruptive behaviour’ or ‘rules’ or ‘safety’ or ‘inclusive’ or ‘teaching practices’ or ‘involvement’ and ‘mechanisms’ or ‘theory’ or ‘theorisation’ or ‘conceptual’ or conceptualisation’ or ‘concept’ or ‘mediators’ or ‘moderators’ or ‘process’ or ‘effects’ or ‘scholarship’ or ‘drivers’ or ‘correlation’ or ‘causation’ or ‘association’ or ‘impact’ or ‘causal pathway’ and ‘low income’ or ‘middle income’ or ‘low and middle income’ or [list of low- and middle-income countries in 2023].

#### Inclusion/exclusion criteria

##### Inclusion

- Study design: Any that provides evidence to help with answering our research questions.
- Documents: LMICs.
- Publication date: any.
- Document type: any document type that will inform the review.
- Population: includes pupils aged 7-16 years.
- Type of report: reporting primary research, a review of research or a theory relevant to the impact of school climate on pupils’ wellbeing in LMICs.
- Setting: conducted in statutory education settings in LMICs.
- Language: English.

##### Exclusion

- Studies reporting only on curricular-based interventions or designed to improve individual knowledge.
- Studies report only on interventions designed to improve the physical infrastructure of the school or classroom.
- Studies containing information on the school climate and wellbeing but not examining the links between the two.
- Studies that do not provide sufficient detail to allow identification of specific aspects of the school climate and wellbeing.
- Not a study of the effects of the school climate on pupils
- Only includes special needs schools.
- Only includes pupils outside the 7-16 years age range.
- In languages other than English.

##### Article screening

*Covidence* will be used to manage article screening and data extraction.

1. Remove duplicates and citations without abstracts or summaries;
2. Two reviewers (PA and RS) will review the titles and abstracts of all retrieved documents captured by our search strategy and code them as ‘potentially relevant’ and ‘not relevant’. Any disagreements will be resolved by discussion or, if necessary, bringing in a third reviewer (LD);
3. Download the full text of potentially relevant documents.

##### Step 3: document appraisal and data extraction

Extract information from the documents, as relevant, that potentially meet our inclusion criteria into an Excel spreadsheet:

1. Document details – title, authors, year of publication, location of study;
2. Country, income group (low, lower-middle, upper-middle) (country income group classified as at the time the research was done);
3. Journal discipline;
4. Aims and objectives of the study;
5. The description – details of the intervention, trainers, design, aim/purpose, length of training;
6. Sample characteristics- age of pupils, sex/gender, socioeconomic status, ethnicity, type of school;
7. The study design and if it is fit for purpose (quality/rigour);
8. The conceptualisation of school climate;
9. Rational for SBCCIs, including any social justice framing;
10. Inner contextual factors (mechanisms) (i.e. the structure, culture and resources of schools that support or inhibit the effectiveness of measures taken to improve the school climate before the intervention was introduced);
11. Outer contextual factors (i.e. the cultural, administrative and policy context within which schools operate that support or inhibit the effectiveness of measures taken to improve the school climate before the intervention was introduced);
12. Proximal outcomes measured;
13. How proximal outcomes were measured – instruments used to measure dimensions of school climate and wellbeing;
14. Student outcomes, behaviour, and experience of interventions;
15. Teacher outcomes, behaviour, and experience of interventions;
16. Agency, stakeholders, including pupils’, teachers’ and parents’ interactions and responses to interventions designed to improve the school climate;
17. Generative mechanisms triggered by the intervention that could have supported change (positive mechanisms), e.g. increased student engagement;
18. Generative mechanisms triggered by the intervention that could have restricted/prevented change (negative mechanisms), e.g. resistance by pupils;
19. Contextual mechanisms that could have supported change (positive mechanisms) commitment of school administration to positive change;
20. Contextual mechanisms that restricted/prevented change (negative mechanisms) ingrained norms and values which support bullying and harassment;
21. Any theoretical explanations identified for explaining the outcomes and the level of the explanation that is, psychological or social;
22. Changes in context following the introduction of SBCIs.

The extraction tool will be piloted. PA, RS, LD, and IS will independently read two documents and complete the extraction table. They will then meet, compare their extraction tables, and agree on necessary modifications.

PA will extract all information. PA will remove any documents containing insufficient relevant data to inform how and why the intervention worked (or did not work) and/or not using credible and trustworthy methods. Documents will be considered relevant if they can help to answer the research questions; that is, they report findings from research on school climate. They will be included as credible if the methods used are adequate for generating the findings; documents will be excluded if they are not based on credible research or are purely anecdotal. The reasons for the exclusion of any document will be noted. A critical realist synthesis does not require that two independent reviewers complete screening for quality or data relevance. However, RS will review any documents PA identifies as not contributing or not using credible and trustworthy methods, with differences being resolved by discussion and, if necessary, by bringing in a third reviewer, LD.

We will provide a descriptive narrative summary of the findings from this stage of the review.

##### Step 5: Analysis and Reporting

The analysis will aim to identify a middle-range interdisciplinary theory that explains how school climate impacts pupils’ wellbeing. It will ‘open the black box’ and identify the mechanisms the interventions triggered that explain how the interventions caused the reported outcomes using the critical realist framework for interdisciplinary research resolution, redescription, retroduction, elimination, identification and refinement (RRREIR) (Table 3) (8,9). An interdisciplinary middle-range theory will then be developed that explains how SBCCIs work, recognising that outcomes will likely differ in different contexts.

**Table 2:**
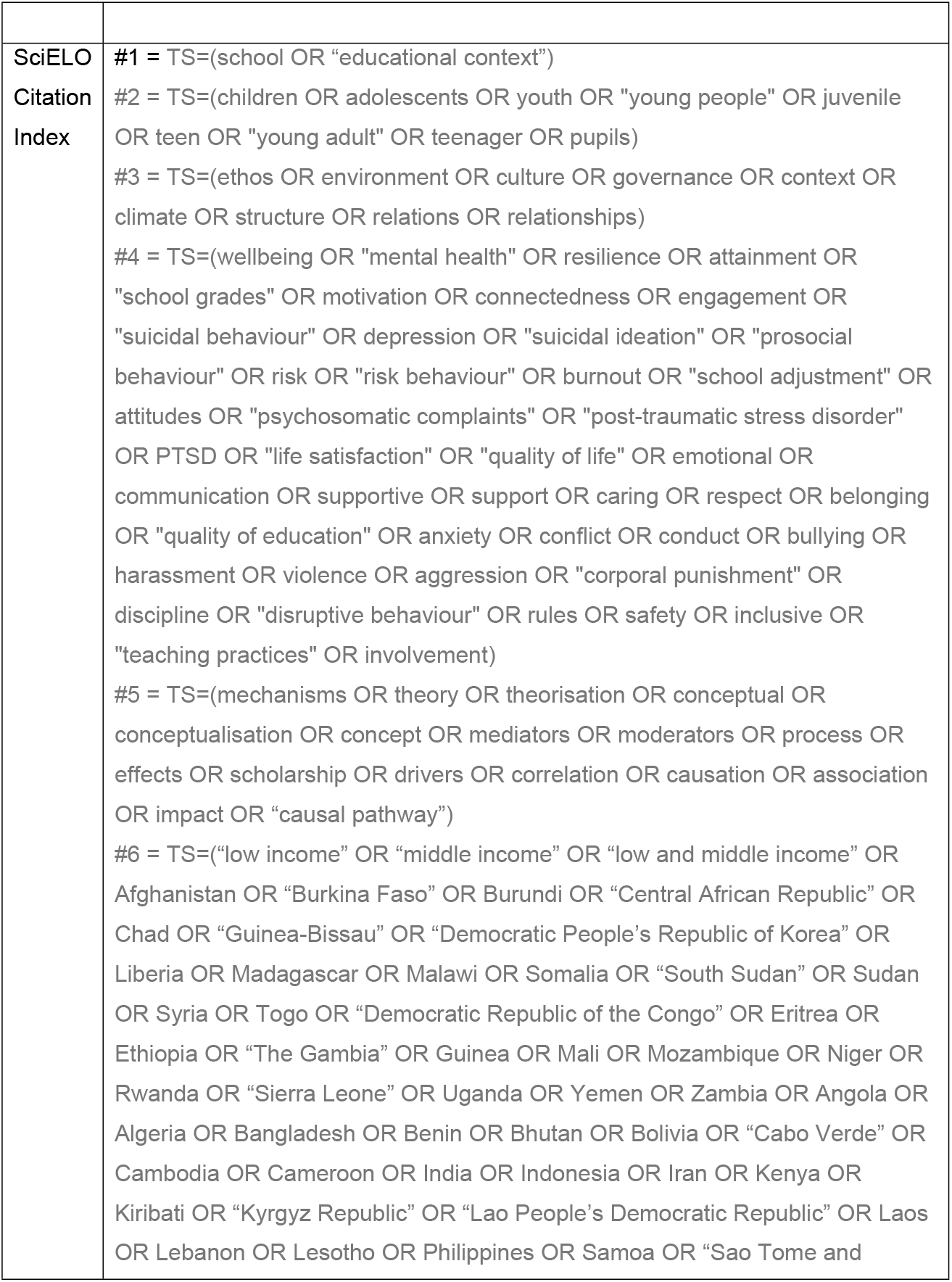
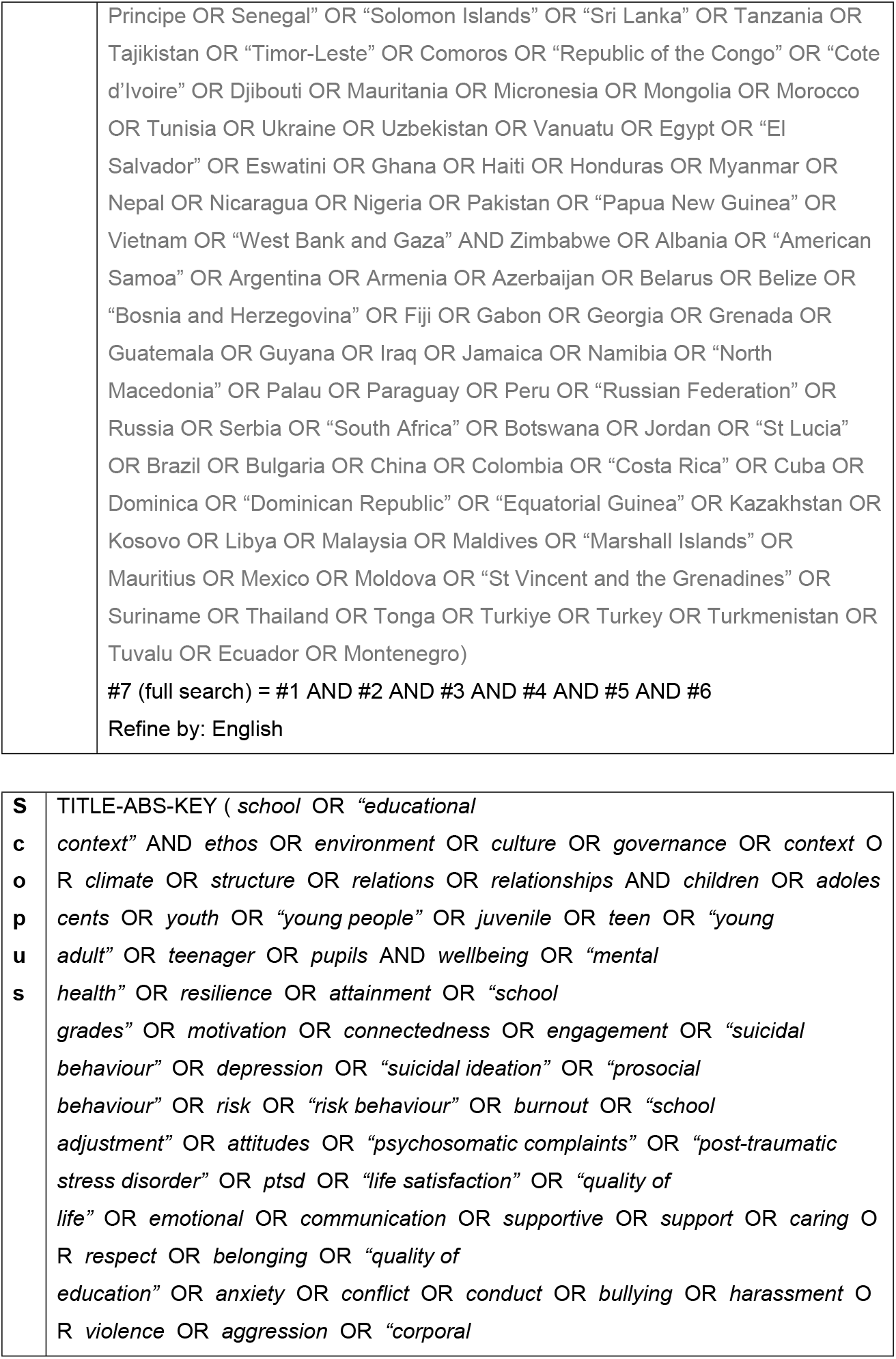
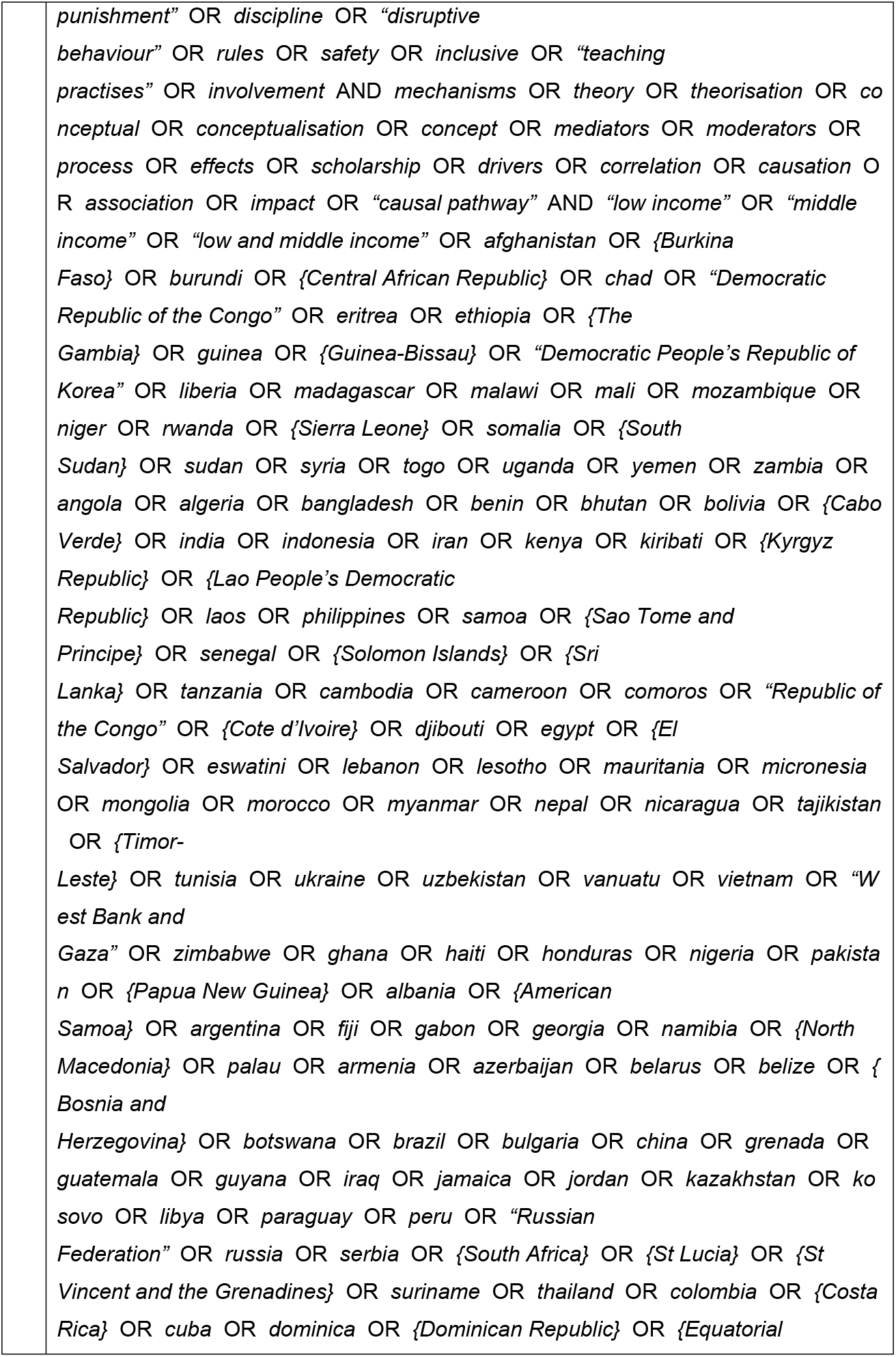
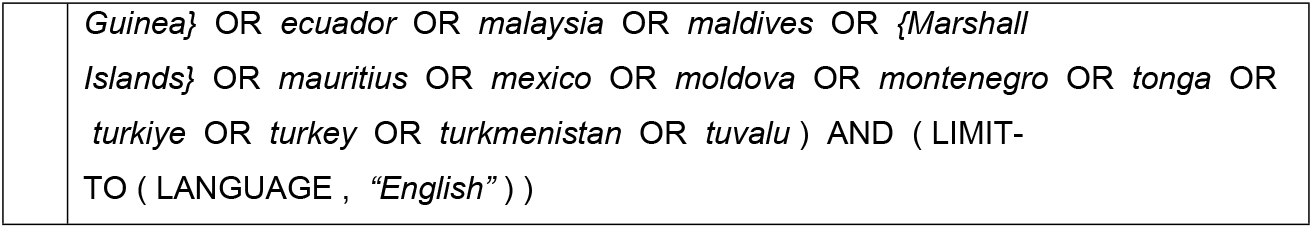
Search Terms for SciELO Citation Index and Scopus

**Table 3:** RRREIc Stages for Interdisciplinary Research

| Planning Phase | Disciplinary Phase | Transdisciplinary Phase |  | Interdisciplinary Phase |  |
| --- | --- | --- | --- | --- | --- |
| Resolution | Redescription | Retrodiction | Elimination | Identification | Refinement |
| Identification of the levels that are included.<br>Biological/<br>Individual<br>Social- Interaction<br>Socioeconomic. | Identifying disciplines and using abduction to develop theoretical explanations for the impact of the intervention on children's mental wellbeing. | Identify how different mechanisms reinforce, moderate, and condition one another to affect the outcome. | Elimination of alternative theoretical explanations. | Identify the most comprehensive interdisciplinary explanation for how school climate promotes children's mental wellbeing. |  |
| Identification of disciplines that research at different levels. | Develop disciplinary theories to explain the impact.<br>Reductionist/ atomistic explanations. | Develop trans-factual theories integrating the disciplinary explanations. | Use judgemental rationality to eliminate theories that lack explanatory power. | Use judgemental rationality to identify the most comprehensive explanation. | Iterative refinement of theory. |

To do this, we will mine the findings from the review to identify the changes that have occurred as a result of the intervention (outcomes), the mechanisms that were triggered by pupils’ (and teachers’) agency and how the context supported or restricted the impact of the interventions CAIMO configurations (40,58) and develop the hypothetical causal links between context, agency, mechanisms and outcomes. To do this, we will use Template Analysis to code the data and identify relevant context, agency, mechanisms and outcomes because it permits using predetermined codes as well as identifying new codes (59). Within each category, findings will be broken down thematically and reported narratively to distinguish between different contexts, agency responses, mechanisms triggered, and outcomes. The key themes that describe processes and causal mechanisms for explaining SBCI outcomes in schools will then be identified. Hypothetical links will then be made between the CAIMO themes, creating potential pathways that account for the impacts of SBCCIs on pupils and why, for whom and under what circumstances these impacts occur.

To account for complexity, it will be necessary to develop non-linear pathways of change showing how the complex interaction of mechanisms (context mechanisms that predate the intervention and those triggered by the intervention) leads to the observed outcomes (43,60– 62). To do this, we will use feedback loop diagrams to model change, showing both mechanisms triggered by the intervention that cause change, those already in the context that supported change (+ve mechanism) and those already in the context and mechanisms triggered by the intervention that restricted or prevented change (-ve mechanisms).

Outcomes will likely be more complex than a dichotomy between morphogenesis (structural change) and morphostasis (structural reproduction) (Table 1; Figuure1).

**Figure 1:**
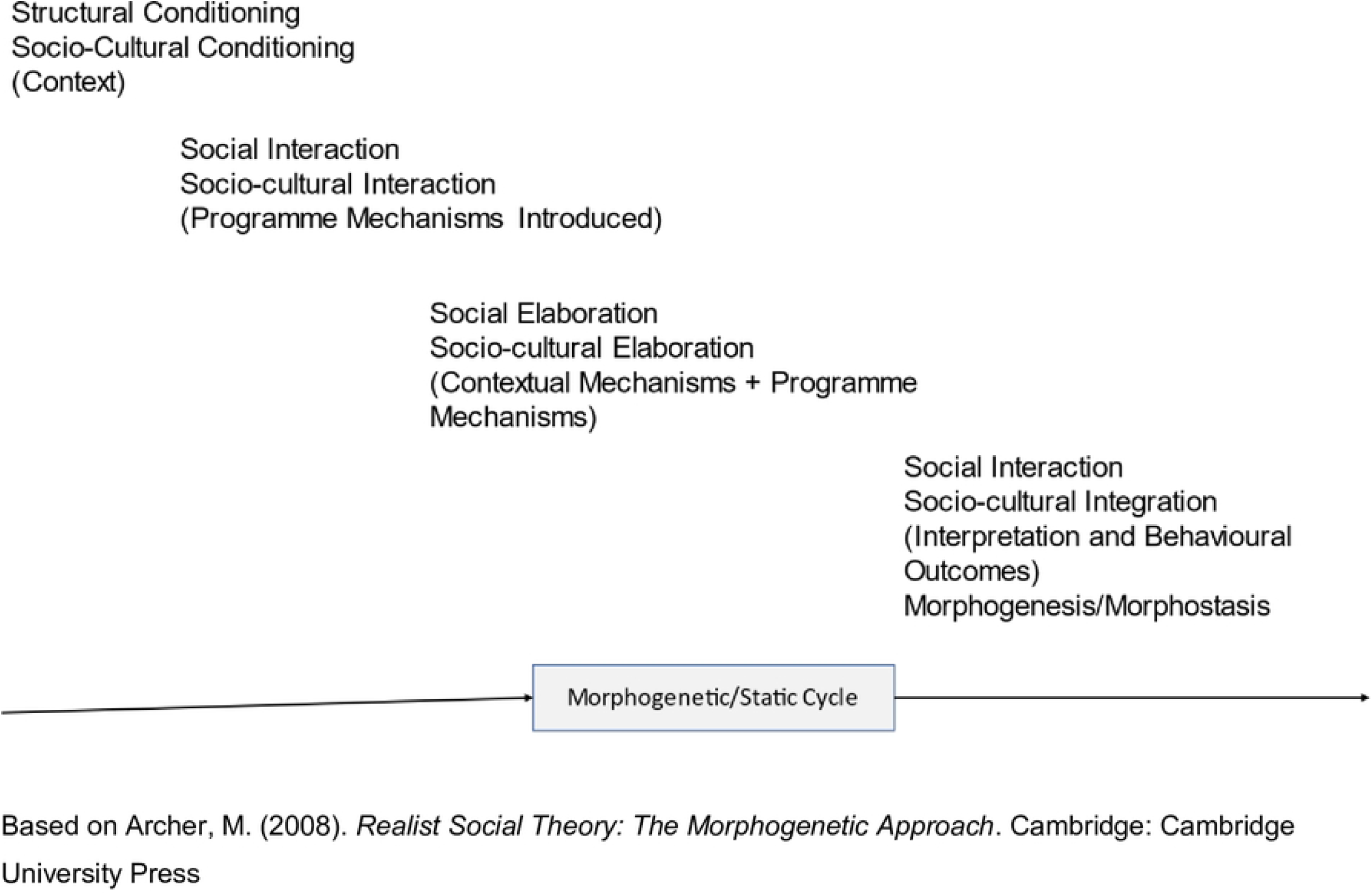
The Morphogenetic Cycle, CAIMO Configuration.

## Ethics

Formal ethics approval is not required for a literature review. However, ethical approval has been obtained from the University of Aberdeen (20^th^ June 2022), Addis Ababa University Ref 111/22/Psy, 21^st^ December 2022), and the University of Rwanda (Ref 03/DRI-CE/012/EN/gi/2023, 25^th^ January 2023) for the research programme, of which this critical realist review forms an integral element.

## Dissemination

We will publish at least one article in a peer review journal reporting the findings from the literature review, conforming to RAMESES publication standards (36) and a policy brief intended for policymakers with the target audience including WHO, UNESCO, UNICEF, the Rwandan and Ethiopian Governments, and the UK and Scottish Governments. Findings from the review will be disseminated via an article in The Conversation, seminar and conference presentations, and podcasts posted on the project website and disseminated by social media.

## Discussion

Our review will be the first critical realist review of the literature on SBCCIs. In the review, we aim to identify the generative mechanisms and social structures that explain how and why SBCCIs promote the wellbeing of pupils. This will enable policymakers and school leaders to understand under what circumstances SBCIs promote pupils’ wellbeing and improve their attainment and for which pupils they work.

The main challenges are likely to be that: there will be little information in the documents on the pre-existing context, and; the documents may not include details of the theoretical reasoning underpinning interventions to enable us to develop middle-range theories.

## Data Availability

N/A

## Ethics approval and consent to participate

Not required for a literature review

## Consent for publication

Not required

## Availability of data and materials

No data was used in preparing this protocol. The data sets that the project generates will be deposited with the UK National Data Archive within six months of the completion of the project under a Creative Commons Attribution 4.0 Unported (CC BY 4.0) license. The training materials produced by the project will be made available under a Creative Commons Attribution 4.0 Unported (CC BY 4.0) license on the project website: https://www.abdn.ac.uk/education/research/cgd/nihr-camw-subsaharan-africa/index.php

## Authors’ contributions

Authors’ Contributions: PA, RS, and LD contributed to the NIHR grant application. PA led the research design and the protocol’s writing and produced the first draft. RS, IS and LD revised drafts and agreed on the final text of this paper.

## Disclaimer

The views expressed in this protocol are the views of the authors alone. They do not necessarily represent the views of the National Institute for Health and Care Research, the UK Government, the Court of the University of Aberdeen, the Board of Directors of the University of Rwanda, or the Board of Addis Ababa University.

## Open access

This is an open-access article distributed in accordance with the Creative Commons Attribution 4.0 Unported (CC BY 4.0) license, which permits others to copy, redistribute, remix, transform and build upon this work for any purpose, provided the original work is properly cited, a link to the licence is given, and an indication of whether changes were made. See: https://creativecommons.org/licenses/by/4.0/.

